# Characterizing and Forecasting Emergency Department Visits Related to COVID-19 Using Chief Complaints and Discharge Diagnoses

**DOI:** 10.1101/2020.06.01.20116772

**Authors:** Phillip Koshute, Rekha Holtry, Richard Wojcik, Wayne Loschen, Sheri Lewis

**Affiliations:** Johns Hopkins University Applied Physics Laboratory, Laurel, MD.

## Abstract

In response to the unprecedented public health challenge posed by the SARS CoV-2 virus (COVID-19) in the United States, we and our colleagues at the Johns Hopkins University Applied Physics Laboratory (JHU/APL) have developed a model of COVID-19 progression using emergency department (ED) visit data from the National Capital Region (NCR). We obtained ED visits counts through targeted queries of the NCR Electronic Surveillance System for the Early Notification of Community-Based Epidemics (ESSENCE). To focus on ED visits by COVID-19 patients, we adjusted the query results for typical ED visit volumes and for reductions in ED volumes due to COVID-19 precautions. With these ED visit data, we fitted a logistic growth model to characterize and forecast the increase in cumulative COVID-19 ED visits. Our model achieves the best fit when we assume that the first NCR visit occurred in early January. We estimate that approximately 15,000 COVID-19 ED visits occurred prior to May 2020 and that approximately 17,000 more visits will occur in subsequent months. We plan to deploy an operational pilot of this model in the NCR ESSENCE environment, assisting local public health authorities as they brace for a second wave of COVID-19. Additionally, we will iteratively assess potential model refinements, aiming to maximize our model’s relevance for local public health authorities’ situational awareness and decision-making.

## 1. Introduction and Background

The spread of the SARS CoV-2 virus (COVID-19) throughout the United States in spring 2020 has posed unprecedented public health challenges^1^. Concerns for shortages in hospital resources have prompted widespread social distancing policies in an attempt to reduce the demand for these scarce resources at any given time. This concept of “flattening the curve” of new daily cases has deep roots within public health practice and was employed as early as the 1918 Spanish Flu Pandemic^2^.

Because these policies are accompanied by strong negative economic and social consequences^3^, it is important to track their actual effect and regularly assess their effectiveness. Foremost, this assessment requires a reliable means of estimating the demand upon public health and hospital resources by actual COVID-19 patients. Once established, these estimates enable the construction of forecast models of the disease’s likely progression within a given population.

Most current forecast models are based on publicly available time series of laboratory test-confirmed cases^4,5^. These data sets are affected, and potentially skewed, by the accuracy of the tests^6^. There are other factors specific to COVID-19 that make laboratory case counts less than ideal: inconsistent clinical criteria for testing^7^; fluctuating availability of tests, which may vary across locations and in time^8^, test processing and reporting delays, and non-standard test reporting (e.g. some states combining antibody and antigen test results^9^). These test-related variations ultimately lead to “apples and oranges” comparisons of differing populations and greatly hinder accurate characterization of COVID-19 progression.

Other current forecast models are based on publicly available time series of deaths attributed to COVID-19^10^. These data sets are similarly affected by criteria that varies by jurisdiction and is applied somewhat subjectively by each death examiner^11^. In some locations, these criteria have also changed over time. Though public awareness of the spread of COVID-19 in the United States surged in early March 2020, recent genomic analysis suggests that disease was likely spreading throughout the country as early as January and February 2020^12^. Consequently, it is unclear how many deaths in January and February are actually attributable to COVID-19.

In contrast to these models based on confirmed cases and attributed deaths, we and our colleagues at the Johns Hopkins University Applied Physics Laboratory (JHU/APL) present the use of emergency department (ED) data to characterize and forecast COVID-19 progression within a metropolitan region. Such data are systematically tracked within the National Capital Region Electronic Surveillance System for the Early Notification of Community-Based Epidemics (NCR ESSENCE) system^13^, which receives data feeds from regional hospitals and urgent care centers.^14^ NCR ESSENCE contains a variety of information from each ED visit at a participating hospital, including chief complaints (CC) and discharge diagnoses (DD). These data are generated as part of routine processes within hospitals and automatically received by ESSENCE. The relevance of various fields, quality factors, and nuances of ED data are well understood, and therefore serves as a valued passive public health disease surveillance data source.

Specific patient groups may be tracked within NCR ESSENCE by defining targeted criteria. For our research, we tracked patients with COVID-Like Illness (CLI) by defining criteria based on CC and DD known to be associated with COVID-19, including fever and shortness of breath. We specifically tracked ED visits in the National Capital Region (NCR), which includes Washington DC, and nearby counties and cities in Virginia and Maryland. We further discuss the resulting data set in Section 2.

Inevitably, these criteria are satisfied by patients that are not actually infected by COVID-19, e.g., those affected by other respiratory illness or by seasonal allergies. We explicitly modeled these “background” patients and subtracted them from raw counts of patients satisfying the CC and DD criteria. Moreover, background patient volumes have likely decreased through intentional efforts to treat patients outside of the ED whenever possible^15^. We also explicitly modeled this effect. We discuss both data processing adjustments in Section 3.

We used the resulting time series to fit a logistic growth model of ED visits by COVID-19 patients. As part of model tuning, we inferred a likely time at which the first COVID-19 case occurred in the NCR. From our model, we forecast progression of ED visits throughout summer 2020. We describe this model in Section 4 and discuss the implications of its results in Section 5. This model is an important step towards establishing COVID-19 forecasting as an operational capability within NCR ESSENCE and providing key insights to local public health officials.

To complement and enhance this model, we suggest avenues for further research in Section 6. Within this section, we also outline our plan for progressing from model development to operational deployment within NCR ESSENCE, a crucial transition for supporting public health authorities’ situational awareness and decision-making. Academic disease modeling researchers often face lengthy timelines before being able to test their models in operational settings or they may lack these opportunities altogether. However, our close ties with public health authorities and the ESSENCE production team uniquely enable us to efficiently assess the operational potential of our model within a dynamic public health surveillance system.

## 2. Data Overview

More than fifteen years ago, ESSENCE was established to provide early outbreak notification in the NCR^16^. In the following years, ESSENCE has been adopted by health systems throughout the United States^17,18^ and further expanded to provide situational awareness and decision support to the public health community^19^.

Public health agencies integrate various data sets using ESSENCE. The primary source of clinical data is ED visit data. The ESSENCE ED “Chief Complaints” field contains mostly pre-diagnostic text strings entered during the hospital patient triage process. The “Discharge Diagnoses” field also contains text strings, as well as some diagnosis codes. To account for the lack of specificity in patient CC and variability in coding among healthcare providers and triage staff, a universal approach is used for syndrome binning. Panels made up of public health and clinical subject matter experts have developed rules-based definitions for assigning patient CC into broad ESSENCE syndromes such as Respiratory, Gastrointestinal, Rash, and Neurological, as well as and subsyndromes such as Influenza, Cough, and Pneumonia.

To capture new or evolving case definitions, public health practitioners also need to develop disease- or condition-specific categories. ESSENCE allows users to customize combination categories based on free-text CC and DD fields. To support the expanded local and national use of ESSENCE, the Centers for Disease Control and Prevention (CDC) National Syndromic Surveillance Program (NSSP) Community of Practice (CoP) brings together public health participants to routinely exchange information on new categories, and to share and refine queries based on disease definitions^20^. At the outset of the COVID-19 crisis, NSSP CoP partners collaboratively developed and refined categories to capture COVID-like-illness (CLI) in ED patient visits.

Seeking to track ED visits by patients infected by COVID-19, we defined criteria for filtering patients based on ESSENCE Subsyndromes and CC and DD categories. Our query searched for all patients with fever, cough, shortness of breath, or any term or codes indicating COVID-19. It combined previous versions of CC and DD categories with additional diagnoses codes to increase sensitivity and specificity in capturing CLI. This query was established to allow patients to meet the CLI definition through the DD codes if not already meeting the definition through their CC. Additionally, the query reflects interim CDC guidance^21^ and explicitly excludes all patients with mention of influenza. In ESSENCE syntax, the query is given as *(,^;Fever and Cough-Sob-DiffBr neg Influenza DD v1;^,or,^;CDC Coronavirus-DD v1;^,or,^;CDC Pneumonia CCDD v1;^,),andnot,(,^;CDC Influenza DD v1;^,)*.

We searched for patients satisfying this query within the NCR implementation of ESSENCE. The NCR is comprised of Washington, DC; four counties (Arlington, Fairfax, Loudoun, and Prince William) and four cities (Alexandria, Fairfax, Manassas, and Manassas Park) in Virginia; and two counties in Maryland (Montgomery and Prince George’s). According to recent censuses, these jurisdictions have a combined population of approximately 5.2 million^22^.

We gathered daily counts of NCR patients satisfying these criteria starting on October 27, 2019 and continuing through May 10, 2020. The time series of these raw counts is shown in Figure 1. Across the NCR, the counts range from 100 to 400 per day.

**Figure 1:**
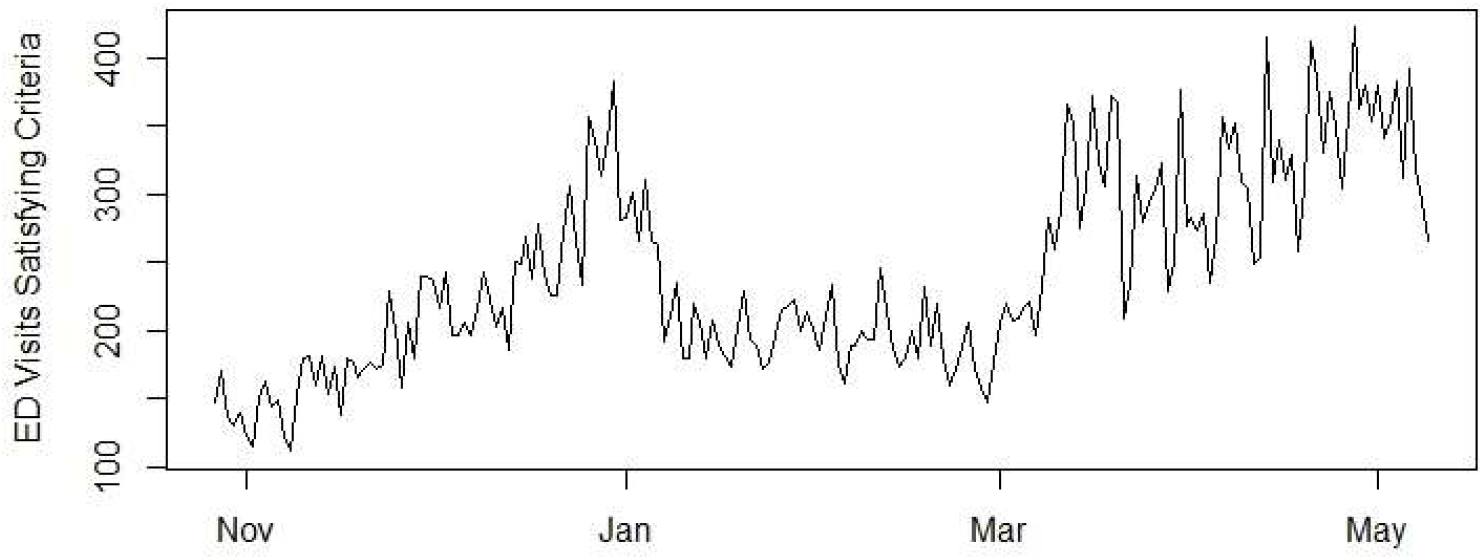
Raw Counts of Emergency Department Visits Satisfying Targeted Criteria.

## 3. Data Processing

Even in the absence of COVID-19, some ED visits satisfy our targeted criteria, as shown in Figure 1. Consequently, it is very likely that not all patients with ED visits satisfying these criteria in recent months have been infected with COVID-19. To estimate the number of ED visits by COVID-19 patients, we must estimate and subtract this set of “background” ED visits. We made this adjustment in two steps: estimating a typical load of background ED visits by week of the year when COVID-19 is not present; and adjusting these estimates to account for the widespread reduction in ED visits by all patients out of precautions related to COVID-19. Sensibly estimating background volume is critical for estimating the number of ED visits by patients actually infected by COVID-19.

To estimate typical background volumes for late 2019 and early 2020, we considered weekly counts of NCR ED patients satisfying the criteria from January 2013 – October 2019. We fitted these values with a normal linear regression model that accounts for seasonal variation and year-to-year increases in ED volumes. Specifically, we modeled each week number as categorical factor and year as a continuous variable. We numbered weeks according to Morbidity and Mortality Weekly Report (MMWR) conventions^23^ and used the lm function in R^24^ with default settings. To calculate typical “expected” volumes for such background ED visits B_w_, we evaluated the resulting model for each week w from October 27, 2019 (week 44) until present. We also considered alternative models for background patient volumes, but ultimately found the normal linear model to be most suitable (cf., Section 5).

The seasonal variation of these counts is illustrated by the fitted model coefficients shown in Figure 2. Larger values for a given week indicate that the volume of ED visits satisfying our targeted criteria is typically larger during that week. The coefficient for Week 1 is 0, i.e., all other weeks’ coefficients are relative to it. Because the targeted ED visits are more likely during Week 1 than in most other weeks, the coefficients for most other weeks are negative. The coefficient for year (with 2020 as year 0) is 1476 and the intercept is 106.

**Figure 2:**
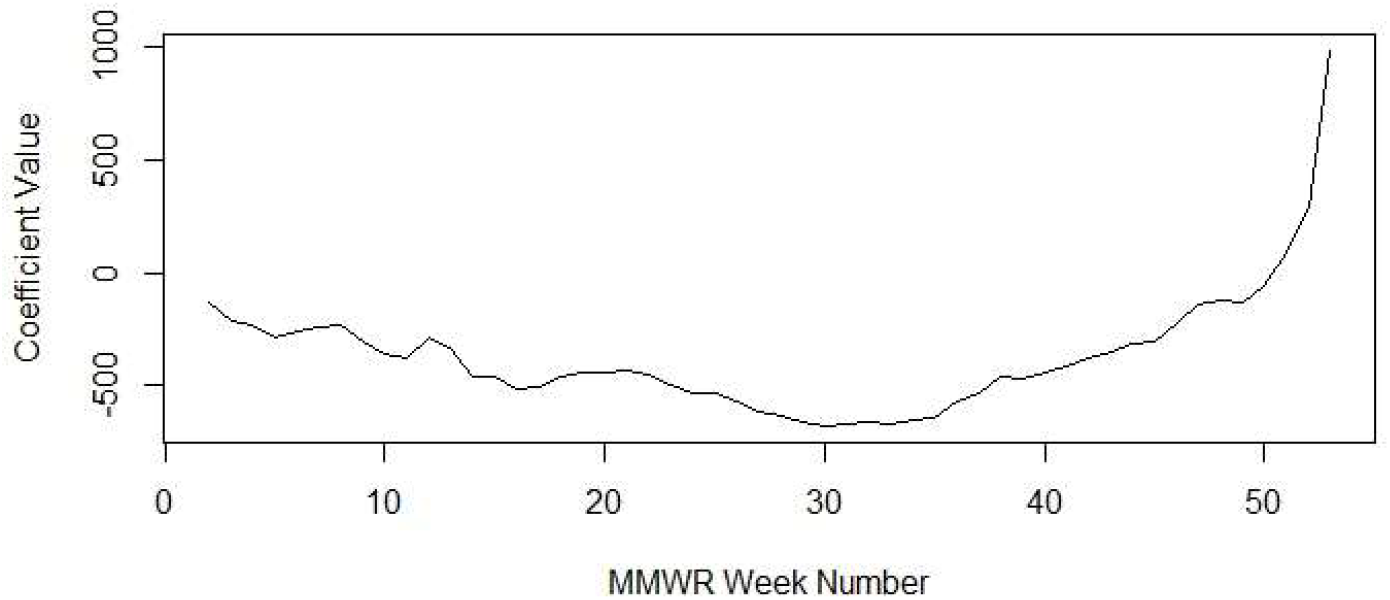
Coefficient Values by Week in Model to Estimate Background Patient Volumes.

As COVID-19 became more widespread, NCR patients became more reluctant to seek ED treatment. For instance, we found that total ED visits in the NCR were nearly 20% lower in April 2020 than in April 2019. We expect that this reluctance to seek ED treatment was also borne by background patients whose ED visits satisfied our target criteria but were not actually infected with COVID-19. Consequently, we reduced our estimates of background patients starting with March 1, 2020 as follows. We tracked ED visits satisfying the “Neurological” syndrome within ESSENCE from October 27, 2019 until present. The Neurological syndrome is characterized by a rules-based definition for assigning patients CC. It consists of a combination of subsyndromes including Altered Mental Status, Confusion, Delirium, and Encephalitis. We selected this syndrome because it typically does not vary seasonally and generally does not overlap with COVID-19 symptoms. We calculated a weekly baseline rate as the mean weekly volume of Neuro patients from October 27, 2019 until February 29, 2020 (i.e., MMWR week 44 in 2019 through MMWR week 9 in 2020). For every subsequent week, we calculated a reduction adjustment factor by dividing that week’s volume by the weekly baseline rate. In March and April 2020, this adjustment factor varies from 0.76 to 0.98. Finally, we multiplied the typical expected background volume in week w by the corresponding adjustment factor a_w_.

To estimate the number of ED visits by patients actually infected by COVID-19 (N_d_) on day d, we calculated the difference between the number of ED visits satisfying our targeted criteria (C_d_) and our adjusted estimates of background ED visits for the corresponding week *w*, i.e.,

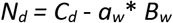

The resulting time series of estimated COVID-19 ED visits is shown in Figure 3. In this plot, negative daily values (e.g., in November) indicate that estimated background counts exceed ED visits satisfying targeted criteria. In order to model cumulative counts of COVID-19 ED visits, we must assume when the first COVID-19 ED visit occurred in the NCR; only estimates to the right of that date are counted as actual COVID-19 ED visits. We discuss our efforts to infer this first-visit date in Section 4. For a given day d, we obtained cumulative counts of COVID-19 ED visits (M_d_) by adding all single-day estimates from the assumed first-visit date until *d*, i.e, 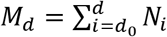.

**Figure 3:**
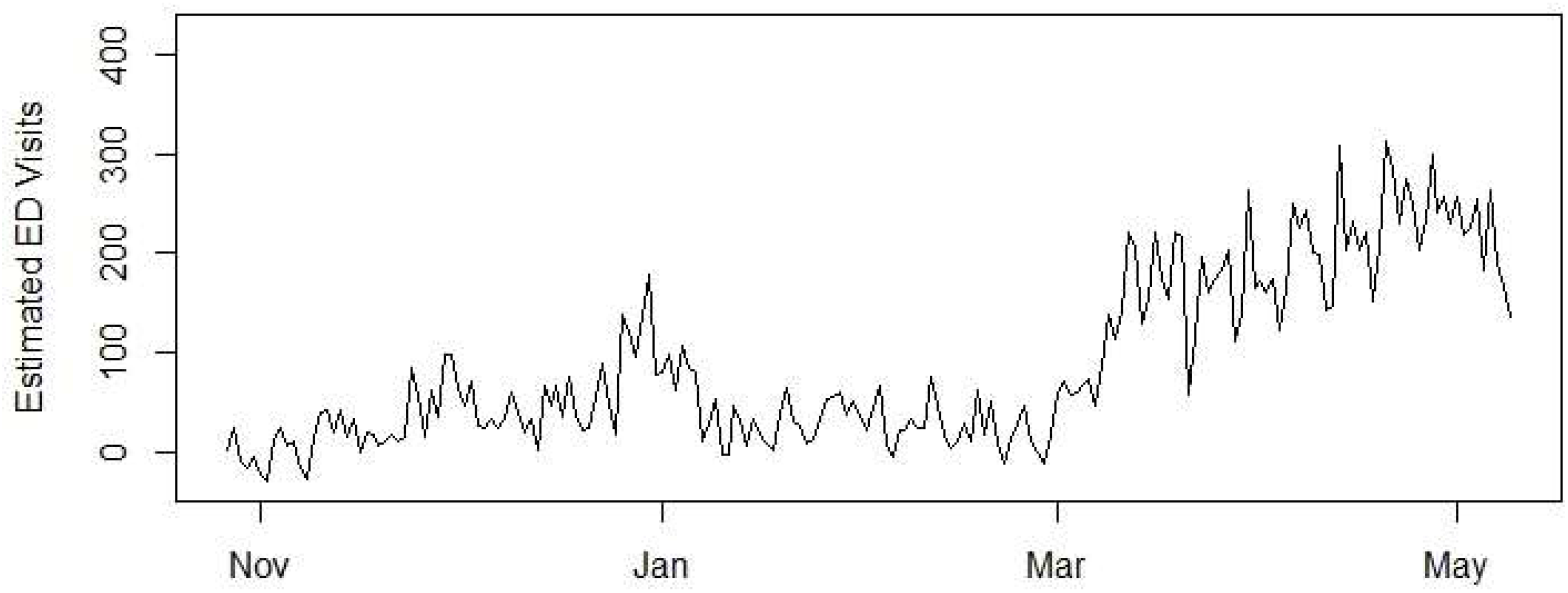
Estimated COVID-19 Emergency Department Visits.

## 4. Model Summary and Results

We fitted cumulative COVID-19 ED visits according to a logistic growth model. Although more sophisticated types of models have been used to model counts of infected patients^25,26^, the logistic growth model is the most straightforward and has been widely used for more than a century^27^. It models each day’s cumulative total according to

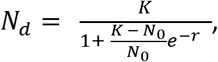

This equation has three parameters: *K* is the maximum number of infected persons (also called “carrying capacity”), *N*_0_ is the initial number of infected persons, and r is the growth rate. To find values for these three parameters and fit the cumulative counts, we used the SummarizeGrowth function in the growthcurver package in R^28^.

As described in Section 3, it is necessary to specify an assumed date for the first NCR COVID-19 ED visit. Determining this date is not straightforward. While the first case of COVID-19 in the NCR was confirmed on March 6, 2020^29^, recent genomic analysis has suggested that the disease may have been present in the region as early as January 2020^30^. This timing aligns with the first confirmed case in the United States in January 2020^31^ and the first known case in France in December 2019^32^. Additionally, this timing is consistent with a comparison of actual counts of ED visits satisfying our targeted criteria in December 2019 and January 2020 with expected background visit counts from that time. As shown in Figure 4, the number of actual visits began to consistently exceed expected counts in late December.

**Figure 4:**
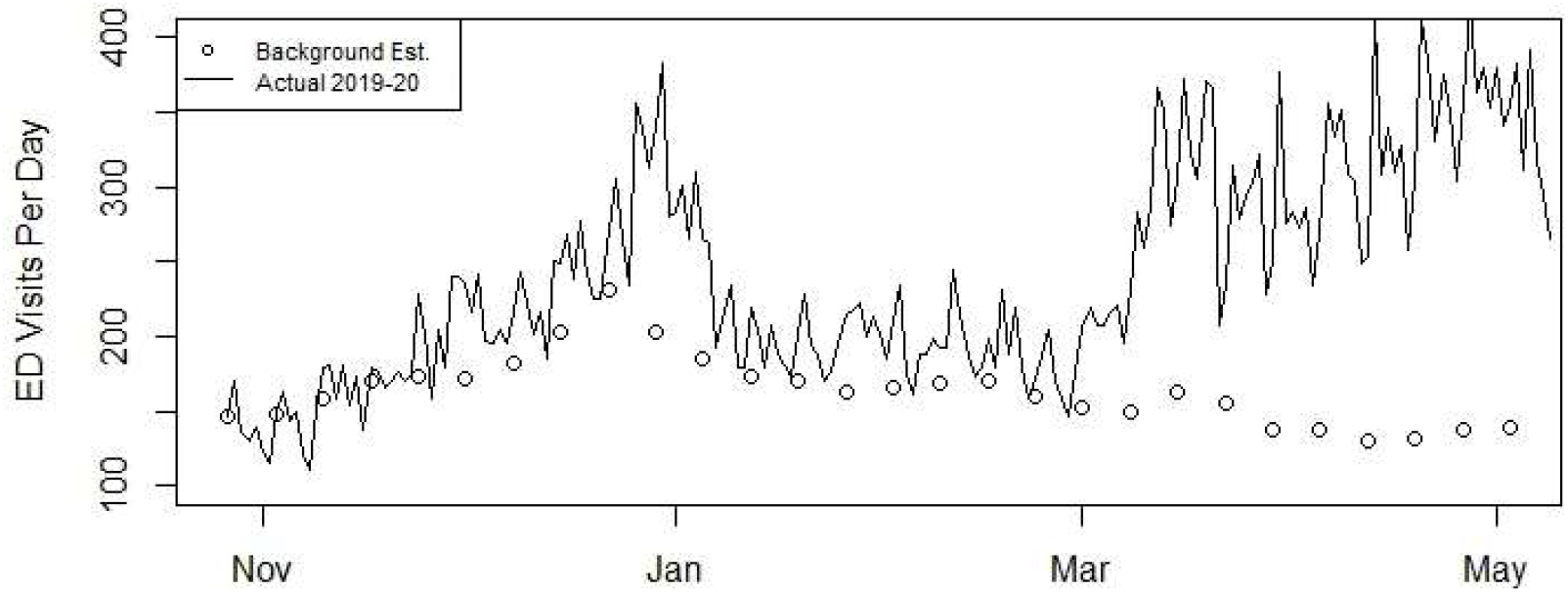
Comparison of Actual and Expected ED Visits Satisfying Targeted Criteria.

Rather than arbitrarily specify an assumed first-visit date, we inferred this date by fitting the model with a range of assumed first-visit dates and assessing which yielded the best model fit. Figure 5 shows the results of these models. For each model, we withheld the most recent days of available counts (April 30 – May 9) and fitted the model only with the available days prior to April 30. This approach enabled us to compare each model’s capability to forecast future days’ counts. In the plots, fitted values of cumulative COVID-19 ED counts for the days that we used to fit the model are shown in blue; withheld counts are shown in green; forecasted values for future days are shown in red; our initial estimates are shown in black. Each model’s forecasting capability can be assessed by comparing the forecasts for withheld days (green line) with the initial estimates for those days (black dots).

**Figure 5:**
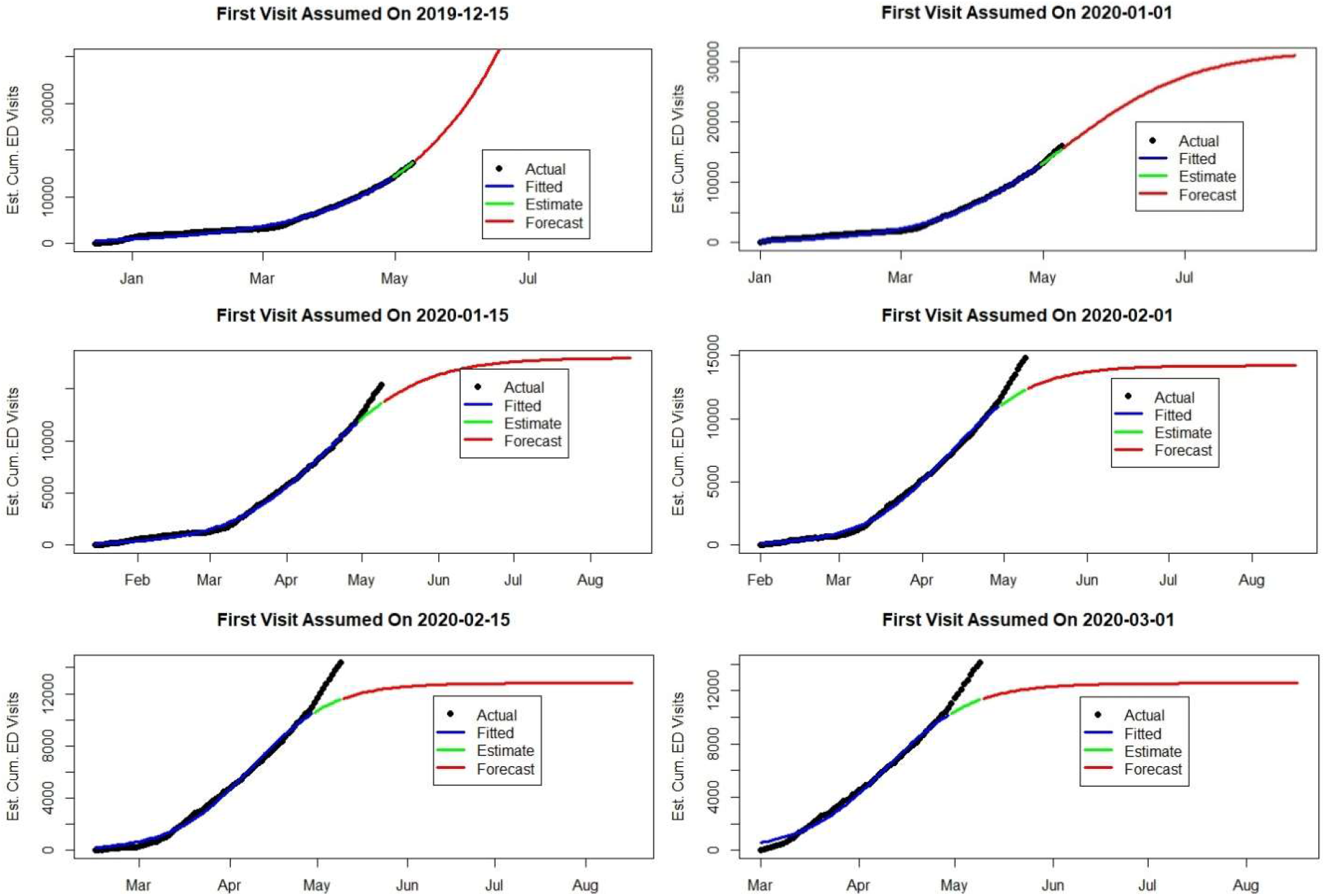
Fitted Models with Different Assumed First-Visit Dates.

We quantified how well each model fits and forecasts the data with three metrics and also validated with a fourth metric. For each model, we calculated the root mean squared error (RMSE) between the initial estimates of cumulative COVID-19 ED visits from the days that we used to train the model and the fitted values for those days, as well as the RMSE between initial estimates from withheld days and the fitted values on those days. For the withheld days, we also considered the day-to-day change in cumulative counts (i.e., the daily counts of visits) for the initial estimates and fitted values, calculating the corresponding RMSE. As further validation, we also considered the fitted value for the eventual maximum number of ED visits. Each of these metrics is shown in Table 1.

**Table 1:** Summary of Model Fit with Different Assumed First-Visit Dates.

| Assumed First-Visit Date | RMSE for Cumulative Counts from Training Days | RMSE for Cumulative Counts from Withheld Days | RMSE for Daily Counts from Withheld Days | Estimated Maximum Number of COVID-19 ED Visits |
| --- | --- | --- | --- | --- |
| 2019-12-15 | 404 | 72 | 58 | $2.16 * 10^{11}$ |
| 2020-01-01 | 258 | 379 | 69 | 32000 |
| 2020-01-15 | 180 | 1187 | 156 | 17900 |
| 2020-02-01 | 188 | 1711 | 210 | 14200 |
| 2020-02-15 | 264 | 1929 | 232 | 13800 |
| 2020-03-01 | 300 | 1908 | 229 | 12500 |

When the first visit is assumed to have occurred on December 15, 2019, the model’s prediction of the eventual total COVID-19 ED visits (over 21 billion) exceeds the total regional population by several orders of magnitude and is clearly too high. Among the other assumed first-visit dates, assuming that the first visit occurred on January 1, 2020 yields the best fit, particularly when forecasting ED visits for days withheld from model training. This model’s forecasts for new counts per day are shown in Figure 6. This model suggests that a peak in COVID-19 visits will occur in early May 2020 and decrease through June and July with approximately 16000 Covid-19 ED visits occurring before and after the peak.

**Figure 6:**
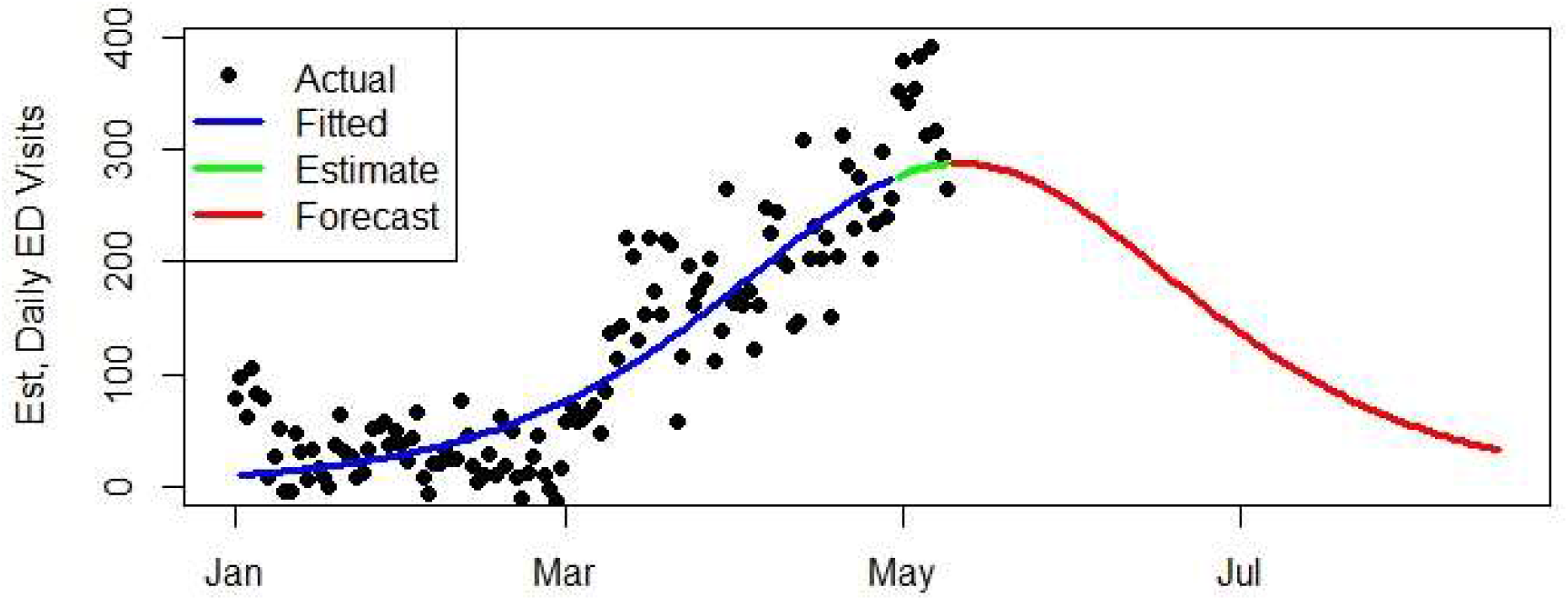
Estimated vs. Fitted Daily Counts of COVID-19 ED Visits, Assuming January 1, 2020 First-Visit.

## 5. Discussion

These results are based on the data sets and modeling assumptions involved in this analysis. We sought to identify all ED visits related to COVID-19 by searching patient’s CC and DD. To compose our query, we focused on the COVID-19 symptoms published by the CDC at the beginning of this analysis, including fever, cough, and shortness of breath. It may be beneficial to repeat this analysis with an updated query including the CDC updated COVID-19 symptoms^33^.

We calculated expected values of weekly background patient volume using a normal linear model. Since the daily number of ED visits are integer counts, it is arguable that Poisson or negative binomial generalized linear models might be more appropriate^34^. However, because the weekly counts are very high (nearly always greater than 500), the corresponding Poisson distribution is very close to a normal distribution. Moreover, while we observed that the overall distribution of weekly counts resembles a Poisson distribution, there is no way to confirm that the distribution of weekly counts by week and month also follow a Poisson distribution since we only have one value for each week-year combination.

To objectively assess what type of model enables the most accurate forecasting, we used data from January 1, 2013 to October 28, 2017 to fit several types of models and compared fitted values with actual counts from October 29, 2017 to October 26, 2019. In addition to normal and Poisson models, we also considered negative binomial models because there is evidence of excessive variance (dispersion) within the counts. For each model, we used the canonical link function. To allow for the possibility of linear relationship contributed by week and year factors, we also constructed models with an identity link function for the Poisson and negative binomial models. The canonical link function for a normal model is the identity link.

As shown in Table 2, the models with identity links provide more accurate fitted values. We ultimately selected a normal linear model for our analysis, since this model is more easily interpreted and more traditional than Poisson or negative binomial models with identity links. Incidentally, for the negative binomial model with identity link, which has the lowest the RMSE, the best fit is also obtained by assuming January 1, 2010 as the first-visit date. In contrast, the best fit for the Poisson model with its canonical link is obtained by assuming a first-visit date in 2019.

**Table 2:** Comparison of Different Model Types for Estimating Expected Background Volumes.

| Model Type | RMSE Between Fitted and Actual Daily ED Visit Counts |
| --- | --- |
| Normal Linear Model | 423 |
| Poisson GLM | 589 |
| Poisson GLM (with Identity Link) | 412 |
| Negative Binomial GLM | 614 |
| Negative Binomial GLM (with Identity Link) | 406 |

Importantly, our model does not incorporate social distancing practices, which has presumably limited the spread of the disease since the onset of related government policies in mid-March 2020 until present. As a result, our forecasts presume that this level of social distancing will continue and are not applicable to scenarios in which these practiced are relaxed.

Our logistic growth model presumes that the spread of the disease is ultimately limited by a majority of the surviving population becoming immune, as illustrated by its underlying Susceptible-Infected-Recovered (SIR) differential equation^35^. However, the overall NCR population of approximately 5.2 million is several orders of magnitude higher than current cumulative totals of COVID-19 ED visits, raising questions of whether other models would be more appropriate. Alternative model types include Gompertz curves^36^ and power law curves^37^, which each assume different progression of the disease throughout the population.

The strong reaction to COVID-19 has aimed to reduce the demand upon hospital resources at any given time. Tracking ED visits related to COVID-19 provides on one aspect of those resources. However, the extent to which these volumes are related to demand upon other resources such as hospital beds, intensive care unit (ICU) beds, and ventilators remains for future analysis.

## 6. Conclusions and Next Steps

As a complement to others’ efforts to forecast COVID-19 progression using publicly available confirmed case data, we have analyzed ESSENCE ED visit data from the NCR. Using these data, we have developed an approach to estimating ED visits by COVID-19 patients and fitted a model that forecasts future volumes of COVID-19 ED visits.

Our model achieves the best fit when the first COVID-19 ED visit is assumed to have occurred in early January 2020. Assuming this first-visit date, the model estimates that approximately 15000 COVID-19 ED visits occurred prior to May 2020 and forecasts that approximately 17000 will occur in the coming months. Although we have focused exclusively on the NCR, this approach could be replicated in other regions, provided that the relevant ED data are available.

As discussed in Section 5, several additional research topics would naturally follow this study, including directly modeling social distancing practices, fitting alternative forecasting models, and identifying the relationship between ED visits and demand for other public health and hospital resources. Such studies are important for continuing to gain insights from these ED data related to the current COVID-19 outbreak, as well as future outbreaks.

Towards maximizing operational relevance for near-term challenges, we plan to deploy a pilot of this model in the NCR ESSENCE environment. This operational capability will assist local public health authorities as they brace for a second wave of COVID-19, which seems likely to coincide with states gradually relaxing social distancing measures. In parallel, we will engage directly with local public health authorities, leveraging their expertise and considering community-specific factors that may impact disease propagation. Through collaboration with healthcare providers and other partners, we will also seek complementary data sets that enable further model validation. Additionally, we will investigate potential benefits of incorporating mobility and viral genomics data. By iteratively assessing these potential model refinements, we aim to maximize our model’s relevance for local public health authorities’ situational awareness and decision-making.

## Data Availability

The data used in this manuscript are not publicly available.

## Acknowledgments

The authors would like to acknowledge the exceptional cooperation of the Maryland Department of Health (MDH), the Virginia Department of Health, (VDH), District of Columbia Department of Health, (DOH), the eight local health departments represented in the NCR Enhanced Surveillance Operating Group (ESOG), and the Metropolitan Washington Council of Governments’ (MWCOG) Health and Medical Regional Programmatic Working Group (H&MRPWG).

ESSENCE Disease Surveillance capability in the National Capital Region is made possible through a grant from the Department of Homeland Security (DHS) Urban Area Security Initiative (UASI); grant number: 18UASI137–01. This work was made possible through a combination of UASI ESSENCE and JHU/APL internal research and development funds. The contents are solely the responsibility of the authors and do not necessarily represent the official views of DHS.

## Footnotes

1 Smith, Nathaniel, and Michael Fraser. “Straining the system: Novel coronavirus (COVID-19) and preparedness for concomitant disasters.” (2020): 648–649.

2 Strochlic, Nina, and Riley D. Champine. “How some cities ‘flattened the curve’during the 1918 flu pandemic’.” National Geographic 27 (2020).

3 Lee, Jong-Wha, and Warwick J. McKibbin. “Estimating the global economic costs of SARS.” Learning from SARS: preparing for the next disease outbreak: workshop summary. Washington, DC: National Academies Press, 2004.

4 Kriston, Levente, and Levente Kriston. “Projection of cumulative coronavirus disease 2019 (COVID-19) case growth with a hierarchical logistic model.” Bull World Health Organ COVID-19 Open Preprints. http://dx.doi.org/10.2471/BLT20 (2020).

5 Petropoulos, Fotios, and Spyros Makridakis. “Forecasting the novel coronavirus COVID-19.” PloS one 15.3 (2020): e0231236.

6 West, Colin P., Victor M. Montori, and Priya Sampathkumar. “COVID-19 testing: the threat of false-negative results.” Mayo Clinic Proceedings. Elsevier, 2020.

7 Weeks, Carly. “Doctors say coronavirus test criteria are inconsistent, could lead to dangerous gaps.” https://www.theglobeandmail.com/canada/article-doctors-say-coronavirus-test-criteria-is-inconsistent-couldlead-to/. Accessed May 15, 2020.

8 Barry-Jester, Anna Maria, et al. “California’s Coronavirus Testing Still A Frustrating Patchwork of Haves and Have-Nots.” https://www.npr.org/sections/health-shots/2020/05/03/849243723/californias-coronavirus-testing-still-afrustrating-patchwork-of-haves-and-have. Accessed May 15, 2020.

9 Young, Stephen “Abbott, Department of State Health Services at Odds Over Conflated Testing Data” https://www.dallasobserver.com/news/texas-coronavirus-testing-conflate-antibodies-11912520. Accessed May 18, 2020.

10 COVID, IHME, and Christopher JL Murray. “Forecasting COVID-19 impact on hospital bed-days, ICU-days, ventilator-days and deaths by US state in the next 4 months.” MedRxiv (2020).

11 Brown, Emma, et al. “Which deaths count toward the covid-19 death toll? It depends on the state.” https://www.washingtonpost.com/investigations/which-deaths-count-toward-the-covid-19-death-toll-it-dependson-the-state/2020/04/16/bca84ae0-7991-11ea-a130-df573469f094_story.html. Accessed May 15, 2020.

12 Baker, Mike and Sheri Fink. “Covid-19 Arrived in Seattle. Where It Went From There Stunned Scientists. https://www.nytimes.com/2020/04/22/us/coronavirus-sequencing.html. Accessed May 15, 2020.

13 Lombardo, Joseph S. “The ESSENCE II disease surveillance test bed for the national capital area.” Johns Hopkins APL Tech Dig 24.4 (2003): 327–334.

14 Holtry, Rekha S., Lang M. Hung, and Sheri H. Lewis. “Utility of the ESSENCE Surveillance System in Monitoring the H1N1 Outbreak.” Online journal of public health informatics 2.3 (2010).

15 Watkins, Chandler. “Hospital ER visits decline amid COVID-19 outbreak.” https://www.ksla.com/2020/04/21/hospital-er-visits-decline-amid-covid-outbreak/. Accessed May 15, 2020.

16 Lewis, Sheri H., et al. “The collaborative experience of creating the National Capital Region Disease Surveillance Network.” Journal of Public Health Management and Practice 17.3 (2011): 248–254.

17 Centers for Disease Control and Prevention. “Assessment of ESSENCE performance for influenza-like illness surveillance after an influenza outbreak--US Air Force Academy, Colorado, 2009.” MMWR. Morbidity and mortality weekly report 60.13 (2011): 406.

18 Wiedeman, Caleb, et al. “Monitoring Out-of-State Patients during a 2017 Hurricane Response using ESSENCE.” Online Journal of Public Health Informatics 10.1 (2018).

19 Rubino, Heather, et al. “Syndromic Surveillance Evaluation of Influenza Activity in At-Risk Sub-Populations.” Online Journal of Public Health Informatics 8.1 (2016).

20 National Syndromic Surveillance Program Community of Practice. Advancing the Science and Practice of Syndromic Surveillance. https://syndromicsurveillance.org/about.html. Accessed May 16, 2020.

21 ICD-10-CM Official Coding Guidelines – Supplement Coding encounters related to COVID-19 Coronavirus Outbreakhttps://www.cdc.gov/nchs/data/icd/ICD-10-CM-Official-Coding-Gudance-Interim-Advice-coronavirus-feb-20-2020.pdf. Accessed May 16, 2020.

22 US Census. “Quick Facts” population tables. https://www.census.gov/quickfacts/fact/table/US/PST045219. Accessed May 8, 2020.

23 Centers for Disease Control and Prevention. “MMWR Weeks.” https://www.n.cdc.gov/nndss/document/MMWR_Week_overview.pdf. Accessed May 15, 2020.

24 R Core Team (2009). R: A language and environment for statistical computing. R Foundation for Statistical Computing, Vienna, Austria. URL https://www.R-project.org.

25 Meyer, Sebastian, and Leonhard Held. “Power-law models for infectious disease spread.” The Annals of Applied Statistics 8.3 (2014): 1612–1639.

26 Mazurek, Jiri, and Zuzana Nenickova. “Predicting the number of total COVID-19 cases and deaths in the USA by the Gompertz curve.” (2020).

27 Verhulst, Pierre-François. “Notice sur la loi que la population suit dans son accroissement.” Corresp. Math. Phys. 10 (1838): 113–126.

28 Sprouffske, Kathleen (2018). Growthcurver: Simple Metrics to Summarize Growth Curves. R package version 0.3.0. https://CRAN-R.projectorg/package=growthcurver.

29 The Office of Governor Larry Hogan. “Governor Hogan Statement Regarding Novel Coronavirus in Maryland.” https://governor.maryland.gov/2020/03/05/governor-hogan-statement-regarding-novel-coronavirus-inmaryland/. Accessed May 15, 2020.

30 Baker, Mike and Sheri Fink. “Covid-19 Arrived in Seattle. Where It Went From There Stunned Scientists. https://www.nytimes.com/2020/04/22/us/coronavirus-sequencing.html. Accessed May 15, 2020.

31 Holshue, Michelle L., et al. “First case of 2019 novel coronavirus in the United States.” New England Journal of Medicine (2020).

32 Deslandes, A., et al. “SARS-COV-2 was already spreading in France in late December 2019.” International Journal of Antimicrobial Agents (2020): 106006.

33 Belluck, Pam. “C.D.C. Adds New Symptoms to Its List of Possible Covid-19 Signs.” https://www.nytimes.com/2020/04/27/health/coronavirus-symptoms-cdc.html. Accessed May 15, 2020.

34 Agresti, Alan. Foundations of linear and generalized linear models. John Wiley & Sons, 2015.

35 Smith, David, and Lang Moore. “The SIR Model for Spread of Disease: The Differential Equation Model.” Loci.(originally Convergence.) https://www.maa.org/press/periodicals/loci/joma/the-sir-model-for-spread-ofdisease-the-differential-equation-model (2004).

36 Mazurek, Jiri, and Zuzana Nenickova. “Predicting the number of total COVID-19 cases and deaths in the USA by the Gompertz curve.” (2020).

37 Meyer, Sebastian, and Leonhard Held. “Power-law models for infectious disease spread.” The Annals of Applied Statistics 8.3 (2014): 1612–1639.

